# Specialist palliative care improves patient experience, reduces bed days and saves money: an economic modelling study of home- and hospital-based care

**DOI:** 10.1101/2025.08.19.25333960

**Authors:** Peter May, Elham Nikram, Therese Johansson, Gemma Clarke, Sarah Mitchell, Irene J. Higginson, Katherine E. Sleeman, Fliss E M Murtagh

## Abstract

**Background:** High-quality evidence suggests that specialist palliative care reduces the odds of dying in hospital. The associated economic implications have not been established.

**Aim:** To evaluate the cost-effectiveness of home- and hospital-based specialist palliative care for adults in England.

**Design:** Health-economic decision-modelling using five-state Markov cohort models with a 24-hour cycle and lifetime horizon.

**Setting/participants:** We evaluated home- and hospital-based care separately for adults in England with poor prognosis. We modelled treatment counterfactuals using Cochrane review evidence of specialist palliative care treatment effects on place of death and quality of life. We estimated place of death distributions, utilisation, unit costs and quality-adjusted life years, and intervention costs, from the literature.

**Results:** Home specialist palliative care was associated with reduced costs of £7,908 per person (95% confidence interval: -18,044 to 395) and increased quality-adjusted life years by 0.035 per person (0.033 to 0.037). Hospital specialist palliative care reduced costs by £6,480 per person (-11,482 to -1,671) and increased quality-adjusted life years by 0.033 per person (0.031 to 0.035). We estimated that for England in 2022, specialist palliative care supported over 20,000 people to die outside of hospital, saved approximately 1.5million hospital bed days and reduced system expenditures by £817million.

**Conclusion:** Specialist palliative care reduces hospital bed days, deaths in hospital and healthcare costs, as well as improving quality of life, among adults in England. A minority who might benefit currently receive specialist palliative care and needs are growing rapidly. Expanding access would likely yield further gains.

**Key statements:** *What is already known about the topic?:*

- Specialist palliative care increases odds of dying outside hospital and improves patient quality of life, but this is a complex intervention and not all who might benefit receive this specialist care.
- Cost-effectiveness of specialist palliative care, and the economic implications of reduced hospital deaths, is a persistent evidence gap for research and policy.

*What this paper adds?:*

- We used decision modelling, a widely-used method in health economics that has not been routinely applied in evaluating palliative care. The key strength of this approach is the capacity to combine data from different sources to estimate cost-effectiveness when there is insufficient trial data to answer the question.
- We found that both hospital-based specialist palliative care and home-based specialist palliative care for adults in England represent excellent value care, reducing the average cost per patient to the NHS while improving patient outcomes.

*Implications for practice, theory or policy:*

- Specialist palliative care is currently accessed by less than half of people who might benefit in England. Expanding access would likely yield further cost-savings and improve outcomes for patients and families.
- Other countries interested in applying these methods to their own data and services can consider using our methodological templates, which we have published open access.

## Introduction

### Background

Less than 1% of people die in high-income countries annually but this group accounts for 8-10% of healthcare spending.^1,2^ Expenditures are primarily driven by acute hospital admissions and often yield poor value with widespread prevalence of potentially modifiable problems including unmanaged symptoms and fragmented care.^3,4^ People with serious medical illness largely prefer to be at home if adequate supports are available,^5^ but a majority in high-income countries visit hospital in the last months of life and hospital is the most common place of death.^3,4,6^

Care of those with serious medical illness in the last months of life is provided by a range of intersecting health and social care providers. Specialist palliative care – defined as care for those with more complex needs which cannot be met by their primary or ‘core’ team alone, requiring a workforce with specialist skills and experience delivering palliative care as their main role^7^ – is one intervention which can potentially to improve outcomes.^8,9^ In England, less than half of people who die from a non-sudden cause get specialist palliative care as they approach end of life.^3^

Economic evidence to inform improvement efforts is thin.^10^ Primary studies in seriously-ill populations are often underpowered for economic evaluation.^10,1112^ There is potential for decision-modelling, combining evidence from a range of sources, to answer economic policy questions where primary data are insufficient,^13,14^ but these have not been widely used to evaluate palliative care.^10^

### Objectives

To estimate the cost-effectiveness of home- and hospital-based specialist palliative care for adults in England.

## Methods

### Study design and intervention

This was an economic modelling study combining data on mortality, place of death, care utilisation, unit costs, quality of life, specialist palliative care receipt and treatment effect estimates. We chose cohort models as appropriate given study resources and data availability.^15^ This study evaluated two models of specialist palliative care: at home, and in acute hospital. We evaluated each model of care separately as insufficient data were available on treatment effects for concurrent receipt of hospital and home specialist palliative care.

Specialist palliative care is delivered by multi-disciplinary teams comprising consultants, staff-grade doctors, specialist nurses, and allied health professionals.^7^ Teams specifically support patients with specialist palliative care needs, including complex symptom management, management of psychosocial concerns, advance care planning, and training and support of the wider workforce in health and social care in providing care for those with serious medical illness. We compared receipt of specialist palliative care to not receiving specialist palliative care, sometimes called usual care. Since treatment effect estimates are drawn from Cochrane reviews, we interpret our main results as reflecting timely, systematic care for an individual.

### Population and setting

People with progressive life-limiting illness may benefit from palliative care at different points in their illness.^7,16^ For adults in England, specialist palliative care is predominantly delivered in the last three to six months of life.^17^ We defined our study population as community-dwelling adults in England, where a clinician would not be surprised if they died in the next three months.^17,18,19^ We distinguished two analytic cohorts: those living at home (and so candidates at the start of the simulation for home/community palliative care), and those admitted to hospital (and so candidates at the start of the simulation for hospital palliative care). We excluded those living permanently in care homes as having different characteristics, patterns of care, survival curves and palliative care interactions.^20^ For population estimates on numbers of people and places of death, the most recently identified data came from 2022.^21^

### Model rationale and characteristics

Descriptive data on people with serious illness, including healthcare use and circumstances of death, are collected widely in high-income countries. Cochrane reviews found that both home- and hospital-based models of specialist palliative care significantly reduced the odds of dying in hospital.^8,9^ Interventions that reduce deaths in hospital are by definition also reducing time in hospital near end of life, but cost-effectiveness implications have not been thoroughly explored.

The operating principle of this study is that since we have good evidence on where a population spends time near end of life, we know their place of death, and we have credible estimates of how and to what extent specialist palliative care affects this place of death, then we can combine these data to model patient flow between different places near end of life, and estimate the associated costs and outcomes with and without receipt of timely specialist palliative care. We assumed that specialist palliative care does not affect survival in either direction, and fixed survival across treatment counterfactuals so all differences in outcome reflect different time in different places and never differential survival.

We conducted the modelling in seven steps. All but the first step were undertaken separately for each analytic cohort (see Appendix). First, we conceptualised a cohort Markov model with five states and a 24-hour time cycle (Figure 1). The live states were defined as physical locations that adults with serious illness in England spend time: home, hospital, care home and hospice. Each of these locations has different cost and quality of life implications, and approximately 98% of deaths occur in one of these places.^21^ In the UK context for seriously-ill adults, a hospice is a facility focused on providing specialist palliative care, and a care home is a facility providing residential care with expert nursing support.

**Figure 1.**
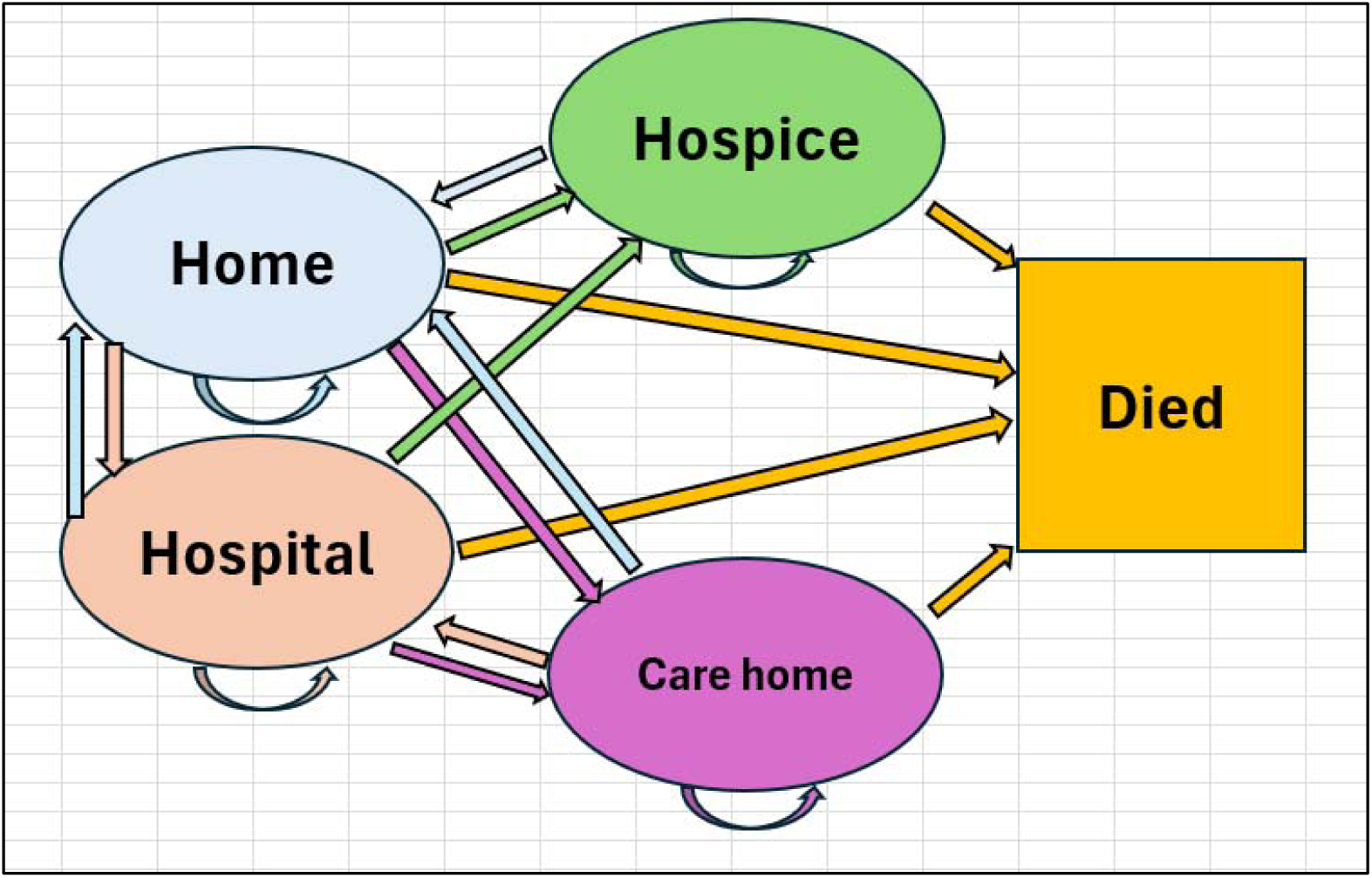
Overview of Markov model structure (24-hour cycle)

Second, we identified relevant cost and outcome parameters for each model state. Third, we specified survival curves. Fourth, we estimated place of death distribution, both for all relevant adults and then stratified by receipt of specialist palliative care. Fifth, we estimated admissions to hospital, care home and hospice, and bed days spent in each of these places, taking into account both survival and proximity-to-death effects. Sixth, we developed the models. We estimated transition probabilities for all adults that delivered model outcomes consistent with the place of death and healthcare utilisation estimated in the prior two steps. In estimating counterfactuals, we allowed treatment status to affect only transition probabilities from the place of care, holding all other parts of the model constant. Seventh, we combined model outputs with outcome parameters to derive our results.

### Measurement of quality of life, and valuation of resources and costs

We sought papers measuring health-related quality of life in older people in the UK with palliative care needs in each of the model states, using EuroQoL EQ5D5L.^22^ We estimated quality-adjusted life years (QALYs) and quantified treatment effects as incremental cost-effectiveness ratios (ICERs). For resource use in each institutional setting, we combined frequency data with unit costs (UK pounds (£), 2022). To estimate the formal costs associated with being at home, we combined frequency data and unit costs for outpatient care, primary care, and district and community nursing.^1^ We estimated informal care costs using the substitution method for a community-based home care worker.^2^ Where costs were identified for a year other than 2022, we adjusted using the UK Consumer Price Index (Health).^23^

### Treatment effects on quality of life and costs

We modelled two additional mechanisms by which specialist palliative care affects model outcomes compared to usual care only:

- Cost of the intervention in each cohort.^24^
- Health-related quality of life. Cochrane reviews,^8,9^ as well as other reviews of high-quality evidence,^17,25,26^ report improved symptom burden and particularly pain management associated with specialist palliative care.

### Perspective, time horizon and discount rate

In primary analysis, we estimated formal care costs from the payer perspective. In secondary analysis we combined formal costs with informal care costs. We estimated QALYs from the patient perspective in all analyses. We combined effects on costs and QALYs to estimate ICERs, interpreting these in the context of a willingness to pay of £20,000 to £30,000 per QALY. Time horizon was the lifetime of the cohort (>99.5% mortality), with a discount rate of 3.5%.^27^

### Uncertainty and additional analyses

In our primary analyses, we applied the odds ratios of treatment effect on place of death and their 95% confidence intervals,^8,9^ generating estimated time in each place for those receiving specialist palliative care and those not in three scenarios: average treatment effect, lower treatment effect, upper treatment effect. Combining these estimates of time in place with the relevant sets of unit costs and QALY values generates an average, lower and upper limit of the estimated effect on costs and QALYs, and the associated ICER.

In our secondary analyses, we repeated the primary analyses but added unpaid family care to the cost of being at home. This analysis from a broader perspective quantifies the extent to which observed savings for the system are displaced to the household. In sensitivity analyses, we checked robustness of our primary results to purposively identified limitations in our data: the cost of a day in acute hospital, the cost of the intervention itself and uncertain QALY effects.

To quantify the estimated effects at the population level, we combined our primary results with data on population health need and specialist palliative care receipt. Since our primary analysis estimates the effect of specialist palliative care per clinical trials, i.e. care provided in a systematic and timely way from a well-defined baseline, but real-world care provision occurs in a less coherent fashion with some patients receiving timely care and others receiving care very late in the disease trajectory,^17^ we incorporated linear dose effects with a range from our primary analysis results (i.e. timely care) and zero (i.e. too late to have any effect).

## Results

### Study parameters

#### Untreated cost and outcome parameters

Estimated cost and QALY inputs by location/state in any base case or untreated scenario are presented in Table 1.

**Table 1.** Study parameters: cost and QALY inputs, base case, both analytic cohorts.

|  | Home | Hospital | Care Home | Hospice | Rationale & references |
| --- | --- | --- | --- | --- | --- |
| **QALYs**<br>Mean (SD) | 0.53 (0.29) | 0.44 (0.29) | 0.57 (0.33) | 0.49 (0.30) | EQ5D5L (where 1 is full health) from location-specific literature on older people in the UK.<sup>28-31</sup> |
| **Daily costs**<br>Mean (Low-High) | £15 | £691<br>(453-794) | £217 | £481<br>(405-735) | Unit cost of a day in a given place.<sup>24,32-34</sup> 'Low' and 'high' used in sensitivity analyses to cost inputs. |
| **Daily informal costs**<br>(secondary analysis only) | £150 |  |  |  | Six hours of unpaid family care when person is at home; hourly cost for a community-based home care worker using substitution method.<sup>2</sup> |

For underlying data and references, see Appendix.

**Table 2.**
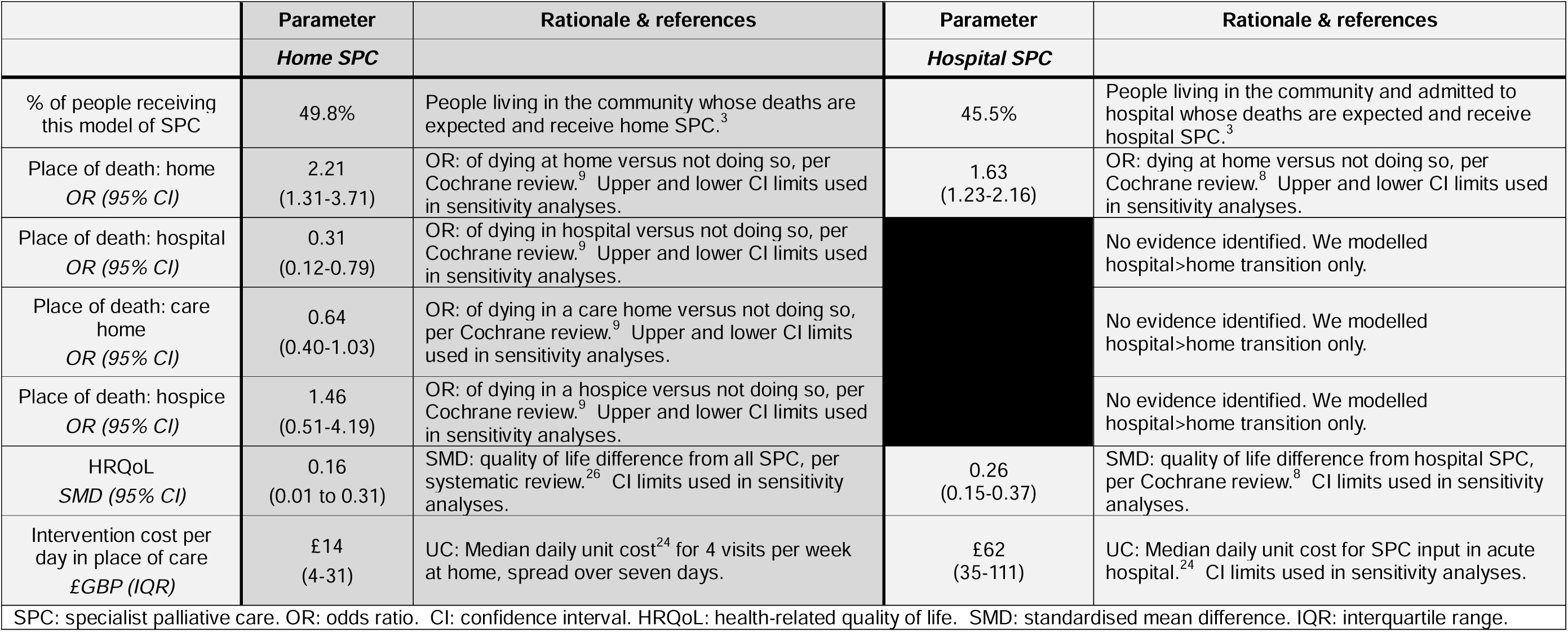
Study parameters: treatment prevalence and effect estimates.

|  | Parameter | Rationale & references | Parameter | Rationale & references |
| --- | --- | --- | --- | --- |
|  | *Home SPC* |  | *Hospital SPC* |  |
| % of people receiving this model of SPC | 49.8% | People living in the community whose deaths are expected and receive home SPC.<sup>3</sup> | 45.5% | People living in the community and admitted to hospital whose deaths are expected and receive hospital SPC.<sup>3</sup> |
| Place of death: home<br>*OR (95% CI)* | 2.21<br>(1.31-3.71) | OR: of dying at home versus not doing so, per Cochrane review.<sup>9</sup> Upper and lower CI limits used in sensitivity analyses. | 1.63<br>(1.23-2.16) | OR: dying at home versus not doing so, per Cochrane review.<sup>8</sup> Upper and lower CI limits used in sensitivity analyses. |
| Place of death: hospital<br>*OR (95% CI)* | 0.31<br>(0.12-0.79) | OR: of dying in hospital versus not doing so, per Cochrane review.<sup>9</sup> Upper and lower CI limits used in sensitivity analyses. |  | No evidence identified. We modelled hospital>home transition only. |
| Place of death: care home<br>*OR (95% CI)* | 0.64<br>(0.40-1.03) | OR: of dying in a care home versus not doing so, per Cochrane review.<sup>9</sup> Upper and lower CI limits used in sensitivity analyses. |  | No evidence identified. We modelled hospital>home transition only. |
| Place of death: hospice<br>*OR (95% CI)* | 1.46<br>(0.51-4.19) | OR: of dying in a hospice versus not doing so, per Cochrane review.<sup>9</sup> Upper and lower CI limits used in sensitivity analyses. |  | No evidence identified. We modelled hospital>home transition only. |
| HRQoL<br>*SMD (95% CI)* | 0.16<br>(0.01 to 0.31) | SMD: quality of life difference from all SPC, per systematic review.<sup>26</sup> CI limits used in sensitivity analyses. | 0.26<br>(0.15-0.37) | SMD: quality of life difference from hospital SPC, per Cochrane review.<sup>8</sup> CI limits used in sensitivity analyses. |
| Intervention cost per day in place of care<br>*£GBP (IQR)* | £14<br>(4-31) | UC: Median daily unit cost<sup>24</sup> for 4 visits per week at home, spread over seven days. | £62<br>(35-111) | UC: Median daily unit cost for SPC input in acute hospital.<sup>24</sup> CI limits used in sensitivity analyses. |
| SPC: specialist palliative care. OR: odds ratio. CI: confidence interval. HRQoL: health-related quality of life. SMD: standardised mean difference. IQR: interquartile range. |  |  |  |  |

#### Counterfactual treatment effect estimates

The parameters for how specialist palliative care interventions changed outcomes are summarised in **Error! Reference source not found.**. Less than half of people living in the community near end of life whose death was expected received home specialist palliative care , and less than half of those admitted to hospital received hospital specialist palliative care. Cochrane review and other meta-analysed trial evidence found that home specialist palliative care increased the odds of dying in own home or hospice, and decreased the odds of dying in hospital or a care home. Hospital specialist palliative care was found to increase odds of dying at home but evidence on dying in other places was not clear. Both models of specialist palliative care were estimated to improve quality of life and we incorporated intervention costs per day in the place where care is provided.

### Summary of main results

The primary analysis results are presented in Table 3. We estimated that, in realising the reduced odds of dying in hospital, total formal costs were £7,908 lower for home palliative care patients than usual care patients (95% CI: -18,044 to 395), while QALYs were higher by 0.035 (95% CI: 0.033 to 0.037). The corresponding ICER was -£227,781 per QALY (95% CI: -482,524 to 12,151). Total formal costs were £6,480 lower for hospital palliative care patients than usual care patients (95% CI: -11,482 to - 1,671), while QALYs were higher by 0.033 (95% CI: 0.031 to 0.035). The corresponding ICERs were negative across the confidence interval, indicating that palliative care dominated usual care in primary analysis.

**Table 3.**
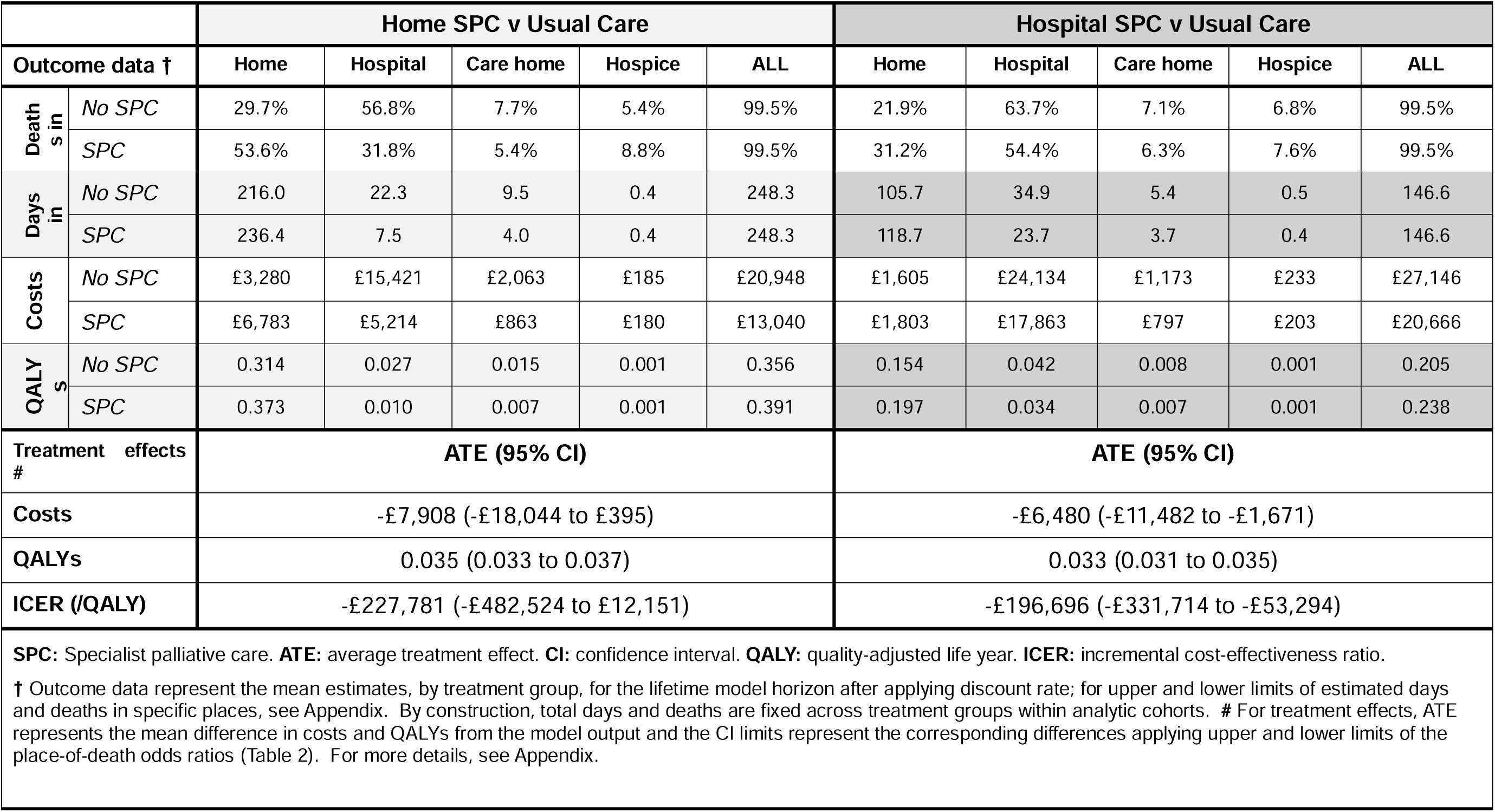
Primary results.

For both models, the estimated cost-savings come from reductions in acute hospital bed days and, to a lesser extent, reduced care home bed days. Increased costs associated with being at home are more than offset by the money saved by reduced use of time in hospital and care home. QALY improvements are driven both by palliative care’s improving effect on outcome, and on the lower QALYs associated with being in hospital compared to being in other places.

### Summary of secondary analysis

The secondary results, where the cost variable incorporates costs from the caregiver perspective, are presented in Table 4. In both cases, the overall estimated effect on costs was smaller than in primary analysis, reflecting how palliative care increases days at home versus hospital, but results were substantively unaffected. At the mean, both models of care dominated the usual care comparator with negative ICERs.

**Table 4.** Secondary analyses: Treatment effects of hospital and home SPC, including informal care costs.

| Home SPC | ATE (95% CI) |
| --- | --- |
| Costs | -£4,840 (-£12,104 to £1,104) |
| ICER (/QALY) | -£139,404 (-£323,675 to £33,933) |
| Hospital SPC | ATE (95% CI) |
| Costs | -£4,520 (-£8,324 to -£863) |
| ICER (/QALY) | -£137,216 (-£240,474 to -£27,517) |
| **SPC:** Specialist palliative care. **ATE:** average treatment effect. **CI:** confidence interval. **QALY:** quality-adjusted life year. **ICER:** incremental cost-effectiveness ratio.<br>Deaths, days and QALY outcomes are unaffected from primary analysis. |  |

### Effect of uncertainty

We reran our primary analyses to test assumptions to the cost of a day in acute hospital; intervention costs; and the co-occurring uncertainty of QoL effects alongside place-of-death effects. Results were substantively unaffected at the mean. See Appendix for further details.

### Population-level estimates

The population-level implications of our results, combining our primary analysis estimates with the number of people estimating to be receiving specialist palliative care, for the calendar year 2022 are presented in Table 5. We estimated that home-based care resulted in 17,098 fewer deaths in hospital (95% CI: -30,681 to -2,985), 1.0million fewer bed days (95% CI: -1.9million to -244,164) and £533.1million lower costs for the NHS (95% CI: - 1.2billion to 31.2million).We estimated that hospital-based care resulted in 3,906 (95% CI: 6,269 to -1,627) fewer deaths in hospital, 491,601 fewer bed days (95% CI: -789,873 to - 205,748) and £283.9million lower costs for the NHS (95% CI: -503.4million to -73.5million).

**Table 5.** Population-level estimates: Treatment effects of home and home SPC on hospital deaths, bed days and healthcare costs in England annually (2022)

| **Home SPC** | **ATE (95% CI)** |
| --- | --- |
| **Hospital deaths** | -17,098 (-30,681 to -2,985) |
| **Hospital bed days** | -1,005,179 (-1,928,774 to -244,164) |
| **Formal costs** | -533,147,283 (-1,220,047,540 to 31,199,212) |
| **Hospital SPC** | **ATE (95% CI)** |
| **Hospital deaths** | -3,906 (-6,269 to -1,627) |
| **Hospital bed days** | -491,601 (-789,873 to -205,748) |
| **Formal costs** | -283,872,364 (-503,411,325 to -73,454,290) |
| **SPC:** Specialist palliative care. **ATE:** average treatment effect. **CI:** confidence interval. |  |

## Discussion

### Key findings

This economic modelling exercise to estimate cost-effectiveness of home- and hospital-based care found that in both cases specialist palliative care represents very good value for patients and decision-makers. Observed cost-effects were realised primarily by reduced days in acute hospital. QALY improvements arose both from palliative care improving this outcome and by higher QALYs associated with places other than hospital. Our main results were robust to sensitivity analyses to selected model parameters. Including informal care costs reduced the estimated cost-effectiveness ratio as caregiver hours increase with time spent at home, but key conclusions were substantively unaffected. Combining our primary analysis with population-level estimates of palliative care need and receipt suggests that each year in England these two models of palliative care support over 20,000 people to die outside of hospital, save approximately 1.5million NHS bed days and reduce system expenditures by £817million.

### What this study adds

The strong causal evidence of effects on place of death and quality of life is long established,^8,9^ but – to the best of our knowledge - this study is the first to quantify the economic implications of these dynamics.^10^ This paper therefore represents the best available evidence of the cost-effectiveness of specialist palliative care for adults in England, and quantifies for the first time the improvements in care and associated cost-savings. At a time when there is widespread unmet need,^35^ and growing needs due to population ageing,^36^ this evidence can inform planning to meet needs, curb cost growth and promote good-value care. Since a minority of people who might benefit currently receive specialist palliative care, it is likely that in the short term expanding access would yield further cost-savings and quality improvements. In the long term, as the population ages, meeting needs will require significant expansion of the specialist workforce. It is also important to consider delivery of palliative and end-of-life care by primary or ‘core’ teams, and how the range of intersecting primary and specialist providers can work most effectively and efficiently together.

The challenges of providing good palliative and end-of-life care during a century of demographic ageing are not unique to England,^37,38^ and other countries can adapt and extend our approach to their own contexts and data. To this end, all modelling materials are provided in the Appendix.

### Strengths and limitations of the study

The strength of this study is that it combines high-quality data from multiple sources to derive credible evidence of cost-effectiveness on interventions that previously have not been subject to widespread economic evaluation. This method harnesses the best available data on specialist palliative care through Cochrane reviews while eliding limitations of other approaches to economic evaluation such as within a clinical trial (when sample size and time horizon are typically insufficient^11^), or observational data (which face challenges of bias^39^ and prospective identification of a sample^40,41^). While confidence intervals in some estimates are large, hospital palliative care dominated the usual care comparator in all scenarios. In the home-based evaluation, even in the most pessimistic scenario from the formal cost perspective, expenditures per patient increased slightly but the corresponding ICER (£12,151 per QALY) well inside the threshold for funding consideration.

The most important limitation is that we used aggregate, population-level data in a cohort model.^15^ There is good evidence that patient quality of life,^42^ care costs,^43^ intervention costs,^24^ patient preferences,^5^ and treatment effects on outcomes^17,44^ all vary by myriad factors including diagnosis, phase of illness, beliefs, household conditions, palliative care dose and supply-side factors. In particular, the timing of palliative care and related dose effects has been shown to be central to cost-effectiveness and clinical efficacy,^45,46^ and we lack a clear relationship between timing and treatment effect estimates to model that more precisely in this study. Variability in timing and components within a given model of care is particularly observable in homecare.^34^

It follows that this study should be the start and not the end of economic analyses that identify how and for whom specialist palliative care makes the biggest difference, addressing *ex ante* specific decision problems for policy, both so that services can optimise allocation of current limited capacity and plan for meeting future needs. Such analyses will optimally use individual-level data and dynamic modelling that allow for more thorough exploration of the demand- and supply-side heterogeneity. While we tested results to key hypothesised parameter uncertainties, we identified other potential uncertainties beyond the scope of our exercise that could be addressed with individual-level data (e.g. survival curves by age and sex; differences in quality-of-life trajectories by location, independent of treatment; unit costs associated with proximity to death or death in an institution; diminishing cost-effectiveness of the intervention as some people prefer not to leave hospital for home).

### Conclusion

This economic modelling study found that specialist palliative care reduces hospital bed days, deaths in hospital and healthcare costs, as well as improving quality of life, among adults in England. A minority who might benefit currently receive it, so expanding access would likely yield further gains. Future research should adapt and extend these findings to other countries and settings, and using individual-level data that will allow for incorporation of demand- and supply-side heterogeneity.

## Supporting information

Appendices: Text and underlying data

## Data Availability

All underlying data are made available as part of this submission.

## Declarations

### Authorship

PM, KES and FEMM conceived and designed the study. PM, TJ, GC and FEMM acquired the data. PM and EN analysed the data. PM, SM, IJH, KES and FEMM interpreted the data. PM drafted the initial paper; all other authors revised the draft critically for important intellectual content and approved the version to be published. All authors have participated sufficiently in the work to take public responsibility for appropriate portions of the content.

### Research ethics and patient consent

This study uses only published literature and aggregate data, and as such ethics approval was not required.

### Data management and sharing

All data and relevant references are provided in the Appendix.

