## Appendices: Text and underlying data for "Specialist palliative care improves patient experience, reduces bed days and saves money: an economic modelling study of home- and hospital-based care": 20250820 PM Appendix.docx

### Overview

Descriptive healthcare data for people with serious illness and poor prognosis are collected widely in high-income countries; in particular, we routinely record formal care activity, e.g. acute hospital admissions, and circumstances of death, including place of death. Cochrane reviews of high-quality evidence on SPC found that both home- and hospital-based models of care significantly reduced the odds of dying in hospital.^1,2^ The operating principle of this study is that if we have good evidence on where our population spends time near end of life, and on their place of death, and we have credible estimates of how and to what extent timely SPC affects this place of death, then we can combine these data to model patient flow between different places near end of life, and estimate the associated costs and outcomes with and without timely receipt of SPC. We assumed that SPC does not affect survival in either direction, and fixed overall survival across treatment counterfactuals so all differences in outcome reflect different time in different places and never differential survival.

We derived distinct analytic cohorts for each model of care, reflecting that the hospital SPC group must by definition be admitted to hospital and so have higher utilisation and higher mortality risk than the home SPC group:

- Community-dwelling (CD) cohort, for the home SPC evaluation
- Hospital admissions (HA) cohort, for the hospital SPC evaluation

We conducted the modelling in seven steps, detailed below and in the spreadsheet ‘Underlying Data’, which is available on Open Science Framework (osf.io/knw9v).^3^ All but the first step were done separately for each analytic cohort.

### Step 1: Model conceptualisation and rationale

Population data on place of death from the Office for National Statistics,^4^ as well as statutory and research data on palliative and end-of-life care specifically,^5,6^ shows that 97% to 98% of deaths in England occur in one of four places: home, hospital, care home, or inpatient hospice. Consequently we devised a model with five states and a 24-hour time cycle (Appendix Figure 1), assuming that deaths occurred only in those places. Of 20 theoretically possible transition probabilities, we modelled 17; we excluded three transitions that, based on data and investigator experience, are rare (though non-zero at the population level): hospice to hospital, and between care home and hospice.

Appendix Figure 1 Conceptual illustration of the model

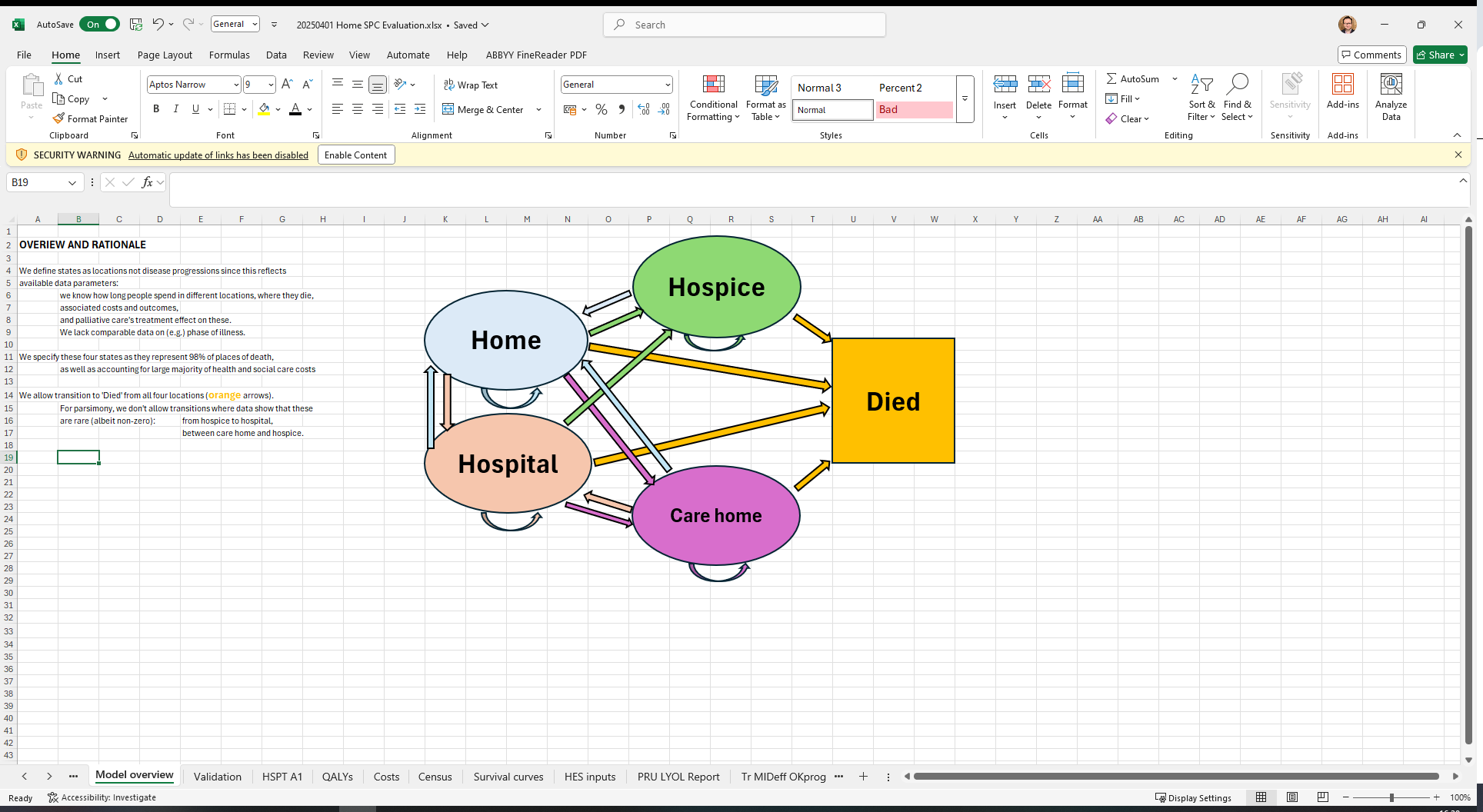

### Step 2: Costs and HRQoL parameters

##### 2a Costs

#### Costs in model states

We estimated the costs of a day at home based on primary and community healthcare use among people approaching end of life, combined with reference costs.^7,8^ We estimated the costs of a day in hospital based on the weighted average of a bed day in an NHS hospital general ward,^9^ adjusting for costs associated with emergency admission and intensive care unit (ICU) stay in a seriously-ill population.^8^ We calculated the weighted average cost of a day in a nursing home across private and local authority providers.^7^ We identified the cost of a day in hospice from the literature.^10^

#### Intervention costs

We estimated the costs of SPC provision based an extensive costing exercise in specialist palliative care in different settings in England.^10^

#### Year of cost data

All costs were adjusted to 2022 using the health component of the consumer price index,^11^ consistent with the population death data.

##### 2b HRQoL

#### HRQoL in model states

We identified studies that reported HRQoL in QALYs for older people living and dying with serious illness in England in each of the four places in the model.^12-15^

#### Treatment effects on HRQoL

For hospital SPC, Bajwah et al synthesise a treatment effect expressed in standardised mean difference (SMD).^1^ For home SPC, Gomes et al report narratively the QoL effects and we did not find a review providing this parameter for home SPC specifically, so we instead used a systematic review and meta analysis of SPC effects that (relative to systematic reviews in the area) has a conservative estimate of effect.^16^

### Step 3: Modelling survival

NICE guidance defines people approaching the end of life as those who are likely to die within 12 months.^17^ People with progressive life-limiting illness may benefit from palliative care at different points in their illness,^18,19^ but in practice for adults in England, SPC is predominantly delivered to people in the last three to six months of life.^20^ A population in (e.g.) the last three, six or 12 months of life can be identified retrospectively and this ‘decedent cohort’ design is widely used for descriptive studies (e.g. the proportion of people to die in a specific location).^21^ However, decedent cohorts have important limitations for evaluative studies because prognosis is prospectively uncertain and implicitly assuming perfect prognostication may bias treatment effect estimates.^22^

There are multiple tools to identify prospectively end-of-life patients in clinical practice or research data, including mortality risk indices,^23^ checklists,^24,25^ weighted algorithmic scoring,^26^ and the ‘surprise question’ (where clinicians are asked, would you be surprised if this patient dies within *t* months?”).^27,28^ Prognostic accuracy varies not only by timeframe but also by age, diagnosis and setting.^27,28^ We defined cohorts according to the positive predictive value (PPV) of prognostication efforts: for a given *n* people who are prospectively identified as approaching end of life, what proportion will die in the next *t* months?

We defined our study population as community-dwelling adults in England, where a clinician would not be surprised if they died in the next three months,^20,29^ leveraging PPV from the ‘surprise question’ literature.^27^ Within this population we distinguished two analytic cohorts: those living at home (and so candidates at the start for the simulation for home/community SPC), and those admitted to hospital (and so candidates at the start for the simulation for hospital SPC). We excluded those living permanently in care homes as having different clinical characteristics, patterns of care, survival curves and palliative care interactions, and as such requiring a separate evaluation.^30^ For population estimates on numbers of people and places of death, the most recently identified official data came from 2022.

In this context, we modelled survival curves based on systematic reviews of the ‘surprise question’ and investigator experience. We modelled separate survival curves for each cohort to reflect differential prognosis by setting. In each case we modelled survival using an exponential distribution, setting the 24-hour mortality rate to deliver a specified 3-month survival rate. SPC models could not affect survival in any scenario.

Appendix Table 1 Survival curve distributions

|  | **CD cohort** | **HA cohort** |
| --- | --- | --- |
| β= | 0.004 | 0.006765 |
| *Survival:* |  |  |
| *3 months* | 69.8% | 54.4% |
| *6 months* | 48.1% | 29.0% |
| *1 year* | 23.2% | 8.5% |
| *2 years* | 5.4% | 0.7% |
| *3 years* | 1.2% | 0.0% |
| *Median days alive* | 173 | 102 |
| *Mean days alive* | 248.3 | 146.6 |

Appendix Figure 2 Survival curves for each cohort

Cumulative survival

Cumlative survival

| Days | Community | Hospital |
| --- | --- | --- |
| 1 | 99.6008% | 99.3258% |
| 2 | 99.2032% | 98.6561% |
| 3 | 98.8072% | 97.991% |
| 4 | 98.4127% | 97.3303% |
| 5 | 98.0199% | 96.6741% |
| 6 | 97.6286% | 96.0223% |
| 7 | 97.2388% | 95.3749% |
| 8 | 96.8507% | 94.7318% |
| 9 | 96.464% | 94.0931% |
| 10 | 96.0789% | 93.4588% |
| 11 | 95.6954% | 92.8286% |
| 12 | 95.3134% | 92.2028% |
| 13 | 94.9329% | 91.5811% |
| 14 | 94.5539% | 90.9637% |
| 15 | 94.1765% | 90.3504% |
| 16 | 93.8005% | 89.7412% |
| 17 | 93.426% | 89.1362% |
| 18 | 93.0531% | 88.5352% |
| 19 | 92.6816% | 87.9383% |
| 20 | 92.3116% | 87.3454% |
| 21 | 91.9431% | 86.7565% |
| 22 | 91.5761% | 86.1716% |
| 23 | 91.2105% | 85.5906% |
| 24 | 90.8464% | 85.0135% |
| 25 | 90.4837% | 84.4403% |
| 26 | 90.1225% | 83.871% |
| 27 | 89.7628% | 83.3056% |
| 28 | 89.4044% | 82.7439% |
| 29 | 89.0475% | 82.186% |
| 30 | 88.692% | 81.6319% |
| 31 | 88.338% | 81.0815% |
| 32 | 87.9853% | 80.5349% |
| 33 | 87.6341% | 79.9919% |
| 34 | 87.2843% | 79.4526% |
| 35 | 86.9358% | 78.9169% |
| 36 | 86.5888% | 78.3848% |
| 37 | 86.2431% | 77.8563% |
| 38 | 85.8988% | 77.3314% |
| 39 | 85.5559% | 76.81% |
| 40 | 85.2144% | 76.2922% |
| 41 | 84.8742% | 75.7778% |
| 42 | 84.5354% | 75.2669% |
| 43 | 84.1979% | 74.7594% |
| 44 | 83.8618% | 74.2554% |
| 45 | 83.527% | 73.7547% |
| 46 | 83.1936% | 73.2575% |
| 47 | 82.8615% | 72.7636% |
| 48 | 82.5307% | 72.273% |
| 49 | 82.2012% | 71.7857% |
| 50 | 81.8731% | 71.3017% |
| 51 | 81.5462% | 70.821% |
| 52 | 81.2207% | 70.3435% |
| 53 | 80.8965% | 69.8692% |
| 54 | 80.5735% | 69.3981% |
| 55 | 80.2519% | 68.9303% |
| 56 | 79.9315% | 68.4655% |
| 57 | 79.6124% | 68.0039% |
| 58 | 79.2946% | 67.5454% |
| 59 | 78.9781% | 67.09% |
| 60 | 78.6628% | 66.6377% |
| 61 | 78.3488% | 66.1884% |
| 62 | 78.036% | 65.7421% |
| 63 | 77.7245% | 65.2989% |
| 64 | 77.4142% | 64.8586% |
| 65 | 77.1052% | 64.4214% |
| 66 | 76.7974% | 63.987% |
| 67 | 76.4908% | 63.5556% |
| 68 | 76.1854% | 63.1271% |
| 69 | 75.8813% | 62.7015% |
| 70 | 75.5784% | 62.2787% |
| 71 | 75.2767% | 61.8589% |
| 72 | 74.9762% | 61.4418% |
| 73 | 74.6769% | 61.0275% |
| 74 | 74.3787% | 60.6161% |
| 75 | 74.0818% | 60.2074% |
| 76 | 73.7861% | 59.8015% |
| 77 | 73.4915% | 59.3983% |
| 78 | 73.1982% | 58.9978% |
| 79 | 72.9059% | 58.6% |
| 80 | 72.6149% | 58.2049% |
| 81 | 72.325% | 57.8125% |
| 82 | 72.0363% | 57.4227% |
| 83 | 71.7487% | 57.0356% |
| 84 | 71.4623% | 56.651% |
| 85 | 71.177% | 56.2691% |
| 86 | 70.8929% | 55.8897% |
| 87 | 70.6099% | 55.5129% |
| 88 | 70.328% | 55.1386% |
| 89 | 70.0473% | 54.7669% |
| 90 | 69.7676% | 54.3976% |
| 91 | 69.4891% | 54.0309% |
| 92 | 69.2117% | 53.6666% |
| 93 | 68.9354% | 53.3047% |
| 94 | 68.6602% | 52.9453% |
| 95 | 68.3861% | 52.5884% |
| 96 | 68.1131% | 52.2338% |
| 97 | 67.8412% | 51.8817% |
| 98 | 67.5704% | 51.5319% |
| 99 | 67.3007% | 51.1844% |
| 100 | 67.032% | 50.8393% |
| 101 | 66.7644% | 50.4966% |
| 102 | 66.4979% | 50.1561% |
| 103 | 66.2324% | 49.8179% |
| 104 | 65.968% | 49.4821% |
| 105 | 65.7047% | 49.1484% |
| 106 | 65.4424% | 48.8171% |
| 107 | 65.1811% | 48.4879% |
| 108 | 64.9209% | 48.161% |
| 109 | 64.6618% | 47.8363% |
| 110 | 64.4036% | 47.5138% |
| 111 | 64.1465% | 47.1935% |
| 112 | 63.8905% | 46.8753% |
| 113 | 63.6354% | 46.5592% |
| 114 | 63.3814% | 46.2453% |
| 115 | 63.1284% | 45.9335% |
| 116 | 62.8764% | 45.6238% |
| 117 | 62.6254% | 45.3162% |
| 118 | 62.3754% | 45.0107% |
| 119 | 62.1263% | 44.7072% |
| 120 | 61.8783% | 44.4058% |
| 121 | 61.6313% | 44.1064% |
| 122 | 61.3853% | 43.809% |
| 123 | 61.1402% | 43.5137% |
| 124 | 60.8962% | 43.2203% |
| 125 | 60.6531% | 42.9289% |
| 126 | 60.4109% | 42.6395% |
| 127 | 60.1698% | 42.352% |
| 128 | 59.9296% | 42.0664% |
| 129 | 59.6903% | 41.7828% |
| 130 | 59.4521% | 41.5011% |
| 131 | 59.2147% | 41.2213% |
| 132 | 58.9783% | 40.9434% |
| 133 | 58.7429% | 40.6673% |
| 134 | 58.5084% | 40.3931% |
| 135 | 58.2748% | 40.1208% |
| 136 | 58.0422% | 39.8503% |
| 137 | 57.8105% | 39.5816% |
| 138 | 57.5797% | 39.3148% |
| 139 | 57.3498% | 39.0497% |
| 140 | 57.1209% | 38.7864% |
| 141 | 56.8929% | 38.5249% |
| 142 | 56.6658% | 38.2652% |
| 143 | 56.4396% | 38.0072% |
| 144 | 56.2142% | 37.7509% |
| 145 | 55.9898% | 37.4964% |
| 146 | 55.7663% | 37.2436% |
| 147 | 55.5437% | 36.9925% |
| 148 | 55.322% | 36.7431% |
| 149 | 55.1011% | 36.4954% |
| 150 | 54.8812% | 36.2493% |
| 151 | 54.6621% | 36.0049% |
| 152 | 54.4439% | 35.7622% |
| 153 | 54.2265% | 35.521% |
| 154 | 54.0101% | 35.2816% |
| 155 | 53.7944% | 35.0437% |
| 156 | 53.5797% | 34.8074% |
| 157 | 53.3658% | 34.5727% |
| 158 | 53.1528% | 34.3396% |
| 159 | 52.9406% | 34.1081% |
| 160 | 52.7292% | 33.8781% |
| 161 | 52.5187% | 33.6497% |
| 162 | 52.3091% | 33.4229% |
| 163 | 52.1003% | 33.1975% |
| 164 | 51.8923% | 32.9737% |
| 165 | 51.6851% | 32.7514% |
| 166 | 51.4788% | 32.5306% |
| 167 | 51.2733% | 32.3112% |
| 168 | 51.0686% | 32.0934% |
| 169 | 50.8648% | 31.877% |
| 170 | 50.6617% | 31.6621% |
| 171 | 50.4595% | 31.4486% |
| 172 | 50.258% | 31.2366% |
| 173 | 50.0574% | 31.026% |
| 174 | 49.8576% | 30.8168% |
| 175 | 49.6585% | 30.609% |
| 176 | 49.4603% | 30.4027% |
| 177 | 49.2628% | 30.1977% |
| 178 | 49.0662% | 29.9941% |
| 179 | 48.8703% | 29.7919% |
| 180 | 48.6752% | 29.591% |
| 181 | 48.4809% | 29.3915% |
| 182 | 48.2874% | 29.1933% |
| 183 | 48.0946% | 28.9965% |
| 184 | 47.9026% | 28.801% |
| 185 | 47.7114% | 28.6068% |
| 186 | 47.5209% | 28.4139% |
| 187 | 47.3312% | 28.2224% |
| 188 | 47.1423% | 28.0321% |
| 189 | 46.9541% | 27.8431% |
| 190 | 46.7666% | 27.6554% |
| 191 | 46.5799% | 27.4689% |
| 192 | 46.394% | 27.2837% |
| 193 | 46.2088% | 27.0998% |
| 194 | 46.0243% | 26.9171% |
| 195 | 45.8406% | 26.7356% |
| 196 | 45.6576% | 26.5553% |
| 197 | 45.4753% | 26.3763% |
| 198 | 45.2938% | 26.1984% |
| 199 | 45.113% | 26.0218% |
| 200 | 44.9329% | 25.8464% |

### Step 4: Primary validation measure: place of death distribution

##### 4a Place of death, treated and untreated pooled

For each analytic cohort we first estimated place of death distribution in the whole population of interest, SPC and no-SPC pooled (see Step 4a in the spreadsheet). We did this by combining ONS data on place of death nationally and survey data on place of death among unexpected deaths,^4,6^ reallocating proportionally the <2% of deaths that occur in some other place to one of the four places in our model.

For home SPC we estimated:

Appendix Table 2 Place of death distribution in CD (home SPC) analytic cohort, treated and untreated pooled

|  | **own home** | **hospital** | **care home** | **hospice** | **all** |
| --- | --- | --- | --- | --- | --- |
| % | 41.7% | 44.6% | 6.6% | 7.1% | 100% |
| n= | 148,584 | 159,093 | 23,556 | 25,368 | 356,601 |

For hospital SPC we estimated:

Appendix Table 3 Place of death distribution in HA (hospital SPC) analytic cohort, treated and untreated pooled

|  | **own home** | **hospital** | **care home** | **hospice** | **all** |
| --- | --- | --- | --- | --- | --- |
| % | 26.3% | 59.6% | 6.8% | 7.3% | 100% |
| n= | 52,456 | 119,089 | 13,570 | 14,582 | 199,696 |

##### 4b/4c Place of death, treated and untreated differentiated

For each analytic cohort we then estimated place of death distribution, differentiating between those receiving SPC and not receiving SPC.

Consider the case of hospital SPC. We estimated place of death with treatment groups pooled above (Appendix Table 3). Bajwah et al.’s Cochrane review estimates that hospital SPC increases odds of dying at home by OR 1.63 (95% CI: 1.23 to 2.16).^1^ Marie Curie survey data shows that of people living in the community (not in a care home) who are admitted to hospital near end of life, 45.5% receive hospital SPC.^6^

These three datapoints provide a set of simultaneous equations that can be solved to estimate place of death by hospital SPC receipt:

Appendix Table 4 Place of death and receipt of hospital SPC, unsolved

| **Die at home?** | **Yes** | **No** | **ALL** |
| --- | --- | --- | --- |
| **Hospital SPC** | A | B | 45.5% |
| **No Hospital SPC** | C | D | 54.5% |
| **ALL** | 26.3% | 73.7% | 100% |

Note: Simultaneous equations: A + B = 45.5; C + D = 54.5; A + C = 26.3; B + D = 73.7; (A/B)/(C/D) = 1.63.

Here is the solution to Appendix Table 4:

Appendix Table 5 Place of death and receipt of hospital SPC, solved for mid-level treatment effect (OR=1.63)

| **Die at home?** | **Yes** | **No** | **ALL** |
| --- | --- | --- | --- |
| **Hospital SPC** | 14.3% | 31.2% | 45.5% |
| **No Hospital SPC** | 12.0% | 42.5% | 54.5% |
| **ALL** | 26.3% | 73.7% | 100% |

And then we reorganise these results within treated/untreated cohorts to establish the outcomes against which counterfactual models can be validated:

Appendix Table 6 Place of death and receipt of hospital SPC, within-cohort outcomes mid-level treatment effect

| **Die at home?** | **Yes** | **No** | **ALL** |
| --- | --- | --- | --- |
| **Hospital SPC** | 31.4% | 68.6% | 100.0% |
| **No Hospital SPC** | 22.0% | 78.0% | 100.0% |

We repeated this process for the lower and upper bounds of Bajwah et al.’s OR confidence interval to derive validation outcomes for weak and strong treatment effects respectively. In the case of hospital SPC, there was no clear evidence on whether the intervention affected deaths in hospices and care homes. We therefore had a choice between (a) fixing the proportion of deaths in those two locations across all scenarios, and (b) validating the model against proportion of deaths at home only and allowing deaths in other places to vary as patients discharged home then moved to other places. Based on discussions among the team, we settled on (b) as more reflective of real-world practice.

For home SPC, Gomes et al provide OR estimates for death in all four places in the model.^2^ We combined these estimates using the same methodology to derive proportion of deaths in each place against which the model can be validated:

Appendix Table 7 Place of death and receipt of home SPC, within-cohort outcomes mid-level treatment effect

| **Place of death** | **own home** | **hospital** | **care home** | **hospice** | **ALL** |
| --- | --- | --- | --- | --- | --- |
| **Home SPC** | 53.7% | 32.1% | 5.5% | 8.8% | 100.0% |
| **No Home SPC** | 29.8% | 57.0% | 7.7% | 5.5% | 100.0% |

We repeated this process for the lower and upper bounds of Gomes et al.’s OR confidence interval to derive validation outcomes for weak and strong treatment effects respectively.

### Step 5: Secondary validation measure: days and admissions

For validating our models of treated and untreated people pooled, we additionally estimated healthcare utilisation that reflect the empirical data. Combining hospital episode statistics data for England,^8^ survey data^6^ and our survival curves, we estimated the mean days in each place in each cohort while controlling for proximity-to-death effects^31^:

Appendix Table 8 Healthcare utilisation, CD cohort (home SPC evaluation)

|  | **Home** | **Hospital** | **Care Home** | **Hospice** | **ALL** |
| --- | --- | --- | --- | --- | --- |
| **Mean days in** | 228.4 | 13.3 | 6.1 | 0.4 | 248.3 |
| **Mean admissions to** | - | 1.23 | 0.23 | 0.09 | - |

Appendix Table 9 Healthcare utilisation, HA cohort (hospital SPC evaluation)

|  | **Home** | **Hospital** | **Care Home** | **Hospice** | **ALL** |
| --- | --- | --- | --- | --- | --- |
| **Mean days in** | 112.7 | 29.1 | 4.4 | 0.3 | 146.6 |
| **Mean admissions to** | - | 0.89 | 0.17 | 0.10 | - |

Hospital simulation starts in hospital. Days in hospital includes this stay, admissions includes only subsequent stays.

From these utilisation data we also inferred provisional transition probabilities between places. These probabilities were only provisional because (a) we are working with aggregate data on time in given places, without detailed data on the proportion of specific transitions (e.g. we have above a credible estimate of how often people are admitted to each of the three institutions, but we don’t have data on what proportion of these admissions occur *from* different places); (b) anchoring all models to place of death while keeping survival fixed means that no single set of transition probabilities suffices across the survival distribution; and (c) each simulation starts in the place of care (home for CD (home SPC); hospital for HA (hospital SPC)).

We finalised these transition probabilities in Step 6 according to valid outcomes of survival, place of death, and admissions to and time in different places.

### Step 6: Populate and validate the models

##### 6a Treated and untreated pooled

For each analytic cohort, we first validated a model where treated and untreated people were pooled. In the ‘Part 2’ spreadsheet we label these the status quo (SQ) models. We have established in Steps 3, 4 and 5 the outcomes against which the model must be validated: survival, place of death, and healthcare utilisation (admissions to and days in a given place). We revised the provisional transition probabilities estimated in Step 5 so that the model delivered valid outcomes for all four measures. We estimated and applied probabilities in five stages of the simulation, each stage reflecting one quintile of the survival distribution, in order to fix survival outcomes in the model to reflect the distribution. We considered model performance satisfactory if mean survival was <0.05 days from the validation benchmark, place of death percentages were within 0.2% of the validation benchmark, and utilisation outcomes were within 0.2 absolute value.

Here is a summary of key inputs and outcomes for the CD (home SPC) cohort:

Appendix Table 10 Transitions and validation, treated and untreated pooled, CD cohort (home SPC evaluation)

| **Transition probabilities** | | | | | |
| --- | --- | --- | --- | --- | --- |
|  | **Home** | **Hospital** | **Care Home** | **Hospice** | **Died** |
| *Home to* | 0.992 | 0.005 | 0.001 | <0.005 | 0.002 |
| *Hospital to* | 0.059 | 0.907 | 0.001 | <0.005 | 0.033 |
| *Care home to* | 0.022 | 0.001 | 0.966 |  | 0.011 |
| *Hospice to* | 0.027 |  |  | 0.790 | 0.183 |
| **Validation outcomes** | | | | | |
| **Survival** | **56 days** | **128 days** | **229 days** | **403 days** | **1325 days** |
| *Model gives* | 80% | 60% | 40% | 20% | <0.5% |
| *Validation***†** | 80% | 60% | 40% | 20% | <0.5% |
| **Place of death** | **Home** | **Hospital** | **Care Home** | **Hospice** | **ALL** |
| *Model gives* | 41.5% | 44.6% | 6.4% | 7.0% | 99.5% |
| *Validation****** | 41.5% | 44.4% | 6.6% | 7.1% | 99.5% |
| **Mean days in** | **Home** | **Hospital** | **Care Home** | **Hospice** | **ALL** |
| *Model gives* | 228.7 | 13.2 | 6.0 | 0.4 | 248.3 |
| *Validation***§** | 228.5 | 13.3 | 6.1 | 0.4 | 248.3 |
| **Mean admissions to** | | **Hospital** | **Care Home** | **Hospice** | **ALL** |
| *Model gives* |  | 1.24 | 0.21 | 0.08 |  |
| *Validation***§** |  | 1.23 | 0.23 | 0.09 |  |
| ***Transition probabilities:*** calculated in five phases (quintiles of the survival distribution). Here we present the median of the values across the five phases.  ***Source of validation outcomes:* †** Step 3 (survival quintiles); ***** Step 4; **§** Step 5. | | | | | |

We calculated equivalent transition probabilities to deliver the relevant valid outcomes for the HA cohort (hospital SPC evaluation); see ‘Part 2’.

##### 6b/c/d Counterfactual (CF) models

For each analytic cohort, we then estimated six counterfactual (CF) scenarios: treated and untreated under mid, weak and strong treatment effect assumptions where those assumptions align to the average, lower and upper OR effects on place of death (Step 4). These mid, weak and strong effect models are reported in 6b, 6c and 6d of ‘Part 2’ respectively.

For any CF scenario, we adjust from the SQ model only specified transition probabilities from the place of care:

- For the home SPC evaluation, we adjust only transition probabilities *from* home to hospital, to care home and to hospice;
- For the hospital SPC evaluation, we adjust only transition probabilities *from* hospital to home.

All other transition probabilities between live states were held constant between the SQ and CF models. This choice was made based on (i) the weight of Cochrane evidence and (ii) expert clinician input that realised changes in place of death are primarily achieved through changing pathways from the place of care (although other changes are also possible, e.g. hospital SPC may reduce the risk of readmission^32^).

Because probability of death differs by place, by moving people between places we impact survival in the model, which we can’t allow because treatment should not impact survival in our assumptions. Therefore we additionally adjust mortality transitions to keep survival constant across CF scenarios. We do this by applying constant weights across places for a given time period, i.e. assuming that relative risk of death between places across timeframes is constant and that changing risk of death in a given place at a given time reflects changing casemix in who is in these different places.

For any CF scenario, we adjust the specified transition probabilities from the place of care and the survival probabilities in order to deliver place of death outcome distribution consistent with that estimated for the given scenario in Step 4.

### Step 7: Compile model outcomes

We combined our cost and HRQoL parameters (step 2) with model output (step 6) on total survival and time spent in different places under different treatment assumptions. The primary and secondary results for the litetime horizon are reported in the main manuscript.

Sensitivity analyses are detailed in the ‘Appendix Part 2’ spreadsheet and summarised below. Given primary results that found interventions to be dominant or highly cost-effective, we tested sensitivity to those parameters we considered most likely to contradict our primary analyses. In all cases, home SPC is dominant at the mean but in the most pessimistic scenarios no longer appears good-value care (£30,000 per QALY < ICER). Hospital SPC is dominant in all scenarios.

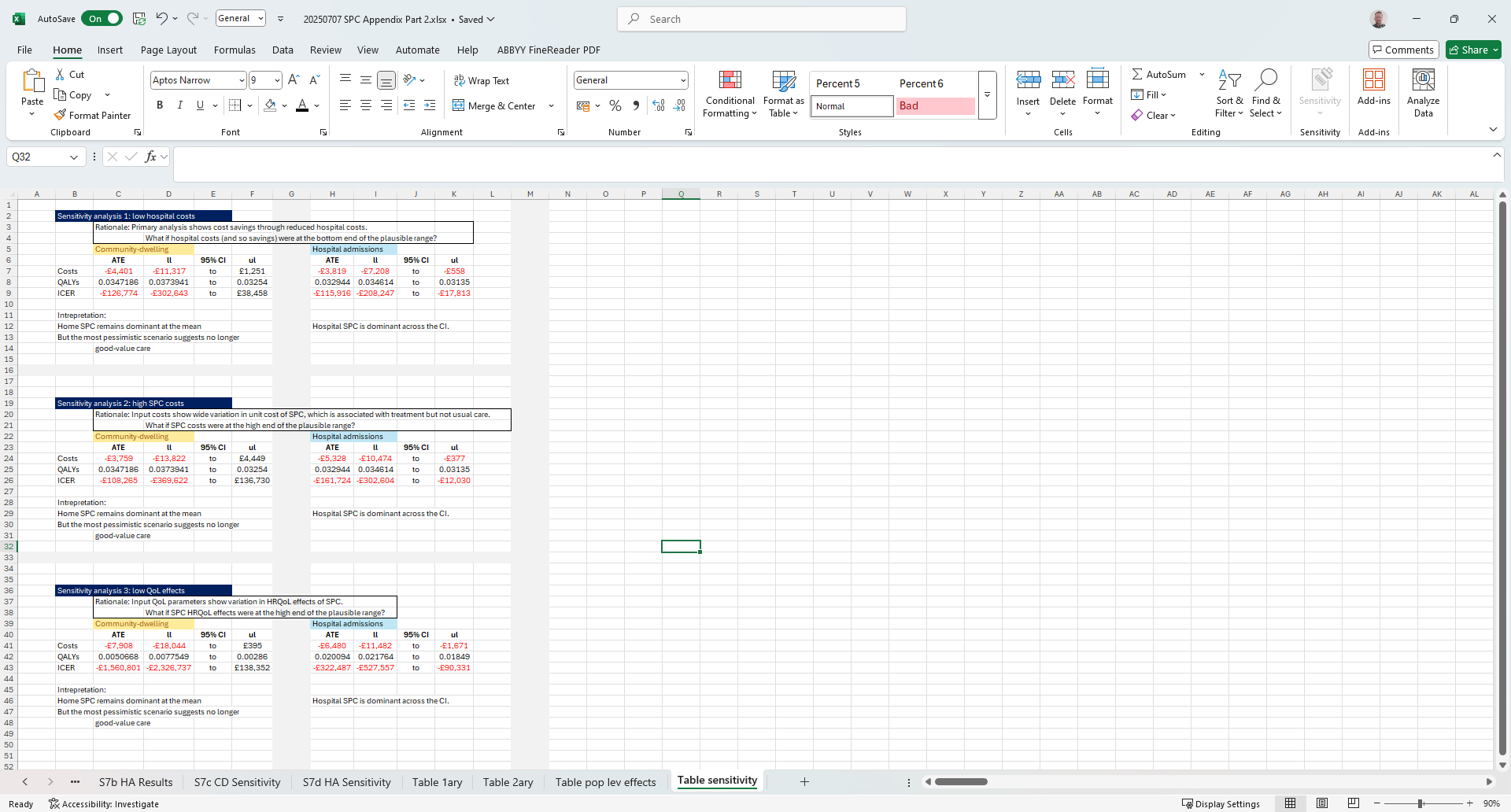

Also reported in the main manuscript are annual estimates at the population level. The calculations for these are detailed in ‘Underlying Data’. Intuitively readers might expect these to reflect our primary effect estimates multiplied by number of patients receiving SPC but this is not the case for two reasons. First, primary results come from a lifetime horizon. A non-negligible minority of patients live more than one year in these models (see Step 3). We extracted cost and QALY effects at 365 days of the simulation for the annual estimates. Second, our primary analysis estimates the effect of SPC per clinical trials, i.e. care provided in a systematic and timely way from a well-defined baseline. Dose effects of SPC are well-documented^33,34^ and real-world care provision occurs in a less coherent fashion with some patients receiving timely care and others receiving care very late in the disease trajectory.^6,20^ Consequently we incorporated an assumption of dose effects: the maximum effect on a given individual was that in our primary analysis (i.e. timely care), the minimum effect was zero (i.e. too late to have any effect), and effects sizes between these extremes were modelled as linear, i.e. the median and mean effect was 50% of the maximum.
